# High-fidelity discrimination of ARDS versus other causes of respiratory failure using natural language processing and iterative machine learning

**DOI:** 10.1101/2021.01.26.21250316

**Authors:** Babak Afshin-Pour, Michael Qiu, Shahrzad Hosseini, Molly Stewart, Jan Horsky, Rachel Aviv, Nasen Zhang, Mangala Narasimhan, John Chelico, Gabriel Musso, Negin Hajizadeh

## Abstract

Despite the high morbidity and mortality associated with Acute Respiratory Distress Syndrome (ARDS), discrimination of ARDS from other causes of acute respiratory failure remains challenging, particularly in the first 24 hours of mechanical ventilation. Delay in ARDS identification prevents lung protective strategies from being initiated and delays clinical trial enrolment and quality improvement interventions. Medical records from 1,263 ICU-admitted, mechanically ventilated patients at Northwell Health were retrospectively examined by a clinical team who assigned each patient a diagnosis of “ARDS” or “non-ARDS” (e.g., pulmonary edema). We then applied an iterative pre-processing and machine learning framework to construct a model that would discriminate ARDS versus non-ARDS, and examined features informative in the patient classification process. Data made available to the model included patient demographics, laboratory test results from before the initiation of mechanical ventilation, and features extracted by natural language processing of radiology reports. The resulting model discriminated well between ARDS and non-ARDS causes of respiratory failure (AUC=0.85, 89% precision at 20% recall), and highlighted features unique among ARDS patients, and among and the subset of ARDS patients who would not recover. Importantly, models built using both clinical notes and laboratory test results out-performed models built using either data source alone, akin to the retrospective clinician-based diagnostic process. This work demonstrates the feasibility of using readily available EHR data to discriminate ARDS patients prospectively in a real-world setting at a critical time in their care and highlights novel patient characteristics indicative of ARDS.

## INTRODUCTION

Characterized by inflammation, hypoxemia, and non-cardiogenic pulmonary edema, the Acute Respiratory Distress Syndrome (ARDS) is described among as many as 10% of all patients admitted to the intensive care unit (ICU)^1^. ARDS is thought to affect over 190,000 patients in the US each year^2^, and the diagnosis of severe ARDS (PaO_2_:FIO_2_ ≤ 100mm Hg) is associated with a nearly 50% mortality rate^1^. Those that survive ARDS experience an elevated risk of cognitive decline and persistent skeletal-muscle weakness^3,4^. Despite the known prevalence of ARDS, due to its potential obfuscation among other disorders requiring ventilation, many clinicians fail to recognize ARDS at the time of respiratory failure^1^ which prevents the initiation of targeted treatments which could improve outcomes (such as prone ventilation, early diuresis and driving pressure-targeted volumes).

The pathophysiologic definition of ARDS has been refined since the disease’s initial description in 2000^5^, but consists of diffuse lung inflammatory changes with increased vascular permeability. The clinical diagnosis is based on bilateral lung infiltrates seen on chest imaging not fully explained by cardiogenic pulmonary edema, and profound hypoxemia^6^. Similar radiologic features and hypoxemia may also be seen in the setting of cardiogenic pulmonary edema (elevated left atrial pressures) making the initial diagnosis challenging in some presentations^7^. As there are no known biomarkers that can diagnose ARDS^8,9^, a real-time implementable patient classification framework that can help discriminate between ARDS versus non-ARDS causes of respiratory failure could assist with earlier identification and potentially reduce the associated morbidity and mortality. It would also allow for prospective enrollment into clinical trials or quality improvement initiatives by assisting first-level automated screening which is particularly helpful for large, multi-hospital clinical studies.

When applied in combination with traditional approaches, machine learning models have shown effectiveness in predicting patient outcomes across disease states, including Sepsis^10^, ICU admission^11^, and Asthma/COPD^12^. Owing to their ability to ingest, integrate, and interpret large volumes of data, ML-based predictive frameworks present substantial potential for the improvement in emergency room operations^13^. Several previous studies have successfully applied ML in the prediction of ARDS (summarized in **Table 1**), demonstrating the utility of this approach. However, while varied in their specific outcomes of interest, none have approached the problem of differentiating ARDS among other causes of respiratory failure requiring mechanical ventilation, which remains a pressing challenge for physicians.

**Table 1.** Recent publications applying ML to predict ARDS onset/diagnosis (limited to non-COVID ARDS studies).

| Study | Design | Setting | N | Dataset | ARDS definition | Ground truth Labeling | Timing of prediction | ML algorithm | Features included | Highly ranked Features | Performance |
| --- | --- | --- | --- | --- | --- | --- | --- | --- | --- | --- | --- |
| Le et al. 2020<sup>23</sup> | Retrospective | ICU | 9919 (296 ARDS) | MIMIC-III | Berlin definition | Berlin definition and mention of bilateral opacities or infiltrates in the patient's radiology report | Detection of ARDS at onset, and prediction at 12-24- and 48 hr prior to onset | XGBoost | Age; Antibiotics; Bilirubin; Blood Culture; Creatinine; Diastolic BP; Fluid Bolus; GCS; HR; INR; Lactate; MAP; Organ Dysfunction; PP; Platelets; Resp. Rate; SpO<sub>2</sub>; Systolic BP; Temp.; Urine Output; WBC; pH | Antibiotics; MAP; PH, RR and SpO<sub>2</sub> (note – this is for the non-mechanically ventilated cohort; data not made available for the mechanically ventilated cohort) | AUC value 0.843 for detection among mechanically ventilated subset |
| Ding et al. 2019<sup>24</sup> | Retrospective analysis of prospectively collected data | ICU | 296 (91 ARDS) | 5 hospitals in Beijing, China | Berlin definition | Automated Berlin Criteria application | Predicting the onset of ARDS 24 after admission | Random forest | minimum respiratory rate, maximum respiratory rate, minimum haematocrit, minimum systolic blood pressure, minimum mean arterial pressure (MAP), maximum heart rate, minimum glucose, minimum white blood cell (WBC) count, minimum heart rate, minimum temperature, minimum sodium level. | Min/max RR; Min hematocrit (ARDS assoc with lower Hct); min SBP; Min MAP; Max HR; Min Glucose (ARDS assoc with higher glucose); Min WBC (ARDS assoc with higher WBC); Min HR; Min Temperature; Min Sodium (ARDS assoc with lower sodium); Age; Min Creatinine; Max MAP; PH; APACHE II | AUC 0.83 |
| Yang et al. 2019<sup>25</sup> | Retrospective | ICU | 8702 | MIMIC-III | Berlin definition | labeled based on Berlin definition P/F < 300, mechanically ventilated, in ICU > 48 hrs | identify ARDS by monitoring P/F values through a variety of noninvasive parameters | XGBoost | SpO<sub>2</sub>; S/F; OSI; PaO<sub>2</sub>; P/F; OI; FiO<sub>2</sub>(%); Temperature (°C); Respiratory rate(b/min); Tidal volume(mL); Tidal volume(mL/kg); Minute ventilation volume(mL/min Peak pressure(cmH<sub>2</sub>O); plateau pressure(cmH<sub>2</sub>O); Mean air pressure(cmH<sub>2</sub>O); PEEP(cmH<sub>2</sub>O); Heart rate(bpm); Nisbp(mmHg); Nidbp (mmHg); Nimbp (mmHg); GCS | SpO<sub>2</sub>; S/F; FiO<sub>2</sub>; PEEP, Mean airway pressure, respiratory rate | AUC 0.9128 |
| Zeiberg et al. 2018<sup>26</sup> | Retrospective | ICU | 1621 (51 ARDS) | Single hospital | Berlin definition | Labeled by two critical care trained physicians | Predicting ARDS from the time of moderate Hypoxia | Regularized logistic regression | baseline patient characteristics (e.g., age, race, and sex) and structured, time-stamped data elements (laboratory values, vital signs, medications administered | Min PaO<sub>2</sub>/FiO<sub>2</sub> ratio; high HR; NI Hgb; High Albumin; Low minimum SpO<sub>2</sub>; : nl Plt count; values decreasing ARDS prediction: missing lactate count; missing PH; Location Schedule Chemotherapy; middle age range (47-58) | AUC of 0.81 (95% CI: 0.73–0.88) |
| Reamaroon et al. 2018<sup>27</sup> | Retrospective | Not reported | 401 (48 ARDS) | Single hospital | Berlin definition | Group of expert clinicians labeled ARDS and non-ARDS based on Berlin definition and onset time between positive patients provided confidence level | Predicting ARDS with uncertain ARDS positive labels | Support Vector Machine | Not reported | Not reported | AUROC of 0.8157 with specificity at 95% sensitivity of 0.5285 on uncertain labels |
| Schenck et al. 2018<sup>25</sup> | Retrospective | Not reported | 4361 ARDS (458 riARDS) | ARDSNet trials | PaO<sub>2</sub>:FIO<sub>2</sub> 2 > 300 on the first study day | Not reported | Prediction of riARDS at Enrollment | Logistic regression | PaO<sub>2</sub>:FIO<sub>2</sub> at screening Change in PaO<sub>2</sub>:FIO<sub>2</sub> from screening to | Not reported | ROC 0.82 (95% CI, 0.78-0.85) |
|  |  |  |  |  | following enrollment; and/or (2) achieving unassisted breathing on the first study day following enrollment and remaining free from assisted breathing for at least 48 h. |  | t |  | enrollment, No use of vasopressor, FIO2 ≤ 0.45 Bilirubin |  |  |
| Zaglam et al. 2014<sup>26</sup> | Retrospective | Pediatric Intensive Care | 90 (53 ARDS) | TARD study: Transfusion Associated Respiratory Distress | presence of bilateral opacities in CXR based Consensus Conference (AECC) criteria | Two intensivists labeled chest radiographs | computer-aided diagnosis system for the detection of ARDS based on CXR | Support Vector Machine on feature extracted from CXR | Spectral features patches and opacities | N/A | Sensitivity of 90.6% at a specificity of 86.5% |
| Chbat et al. 2012<sup>28</sup> | Retrospective | ICU | 526 (216 ALI) | Multidisciplinary Epidemiology and Translational Research in Intensive Care (METRIC) DataMart | ALI: American European Consensus Conference (AECC) criteria | Two independent physician review of radiologic assessment of bilateral infiltrates and PaO2/FiO2 ratio thresholds | Early detection of ALI | Rule-based fuzzy inference systems, Bayesian networks, finite state machines | Pre-ICU data included patients' chronic diseases and surgical history; ICU data included demographics, monitoring data, medications, ventilation settings, laboratory findings, and current health status information. | Not reported | 71.7–92.6% sensitivity and 60.3–78.4% specificity |
| Koenig et al. 2011<sup>29</sup> | Prospective | ICU | 1270 (84 ALI) | Hospital of the University of Pennsylvania | ALI: American European Consensus Conference (AECC) criteria | Screening by research coordinator with two physician adjudication when discrepancy between automated screen detection and coordinator detection | Early recognition of Acute lung injury (ALI) | No ML model. Automated Electronic Acute Lung Injury Screening tool (ASSIST screening tool) | ABG results CXR report | Not reported | Sensitivity of 97.6% (95% confidence interval, 96.8–98.4%) and a specificity of 97.6% (95% confidence interval, 96.8–98.4%) |
| Solti et al. 2009<sup>15</sup> | Prospective | All in-patients | 856 | University of Washington Health System | ALI: American European Consensus Conference (AECC) criteria | 11 Physicians annotated 96 CXR reports as ALI or not | Machine-learning based classification ALI from chest radiography | Maximum Entropy (MaxEnt) algorithm | N/A | N/A | Recall=0.91, Precision=0.90 and F-measure=0.91 |
| Herasevich et al. 2009<sup>30</sup> |  |  | 3795 (325 ALI) | Single Healthcare System | ALI: American European Consensus Conference (AECC) criteria | Two clinician review of 'sniffer' detected cases | Two features: PaOs/FiO2 ratio and radiology report if included 'bilateral' and 'infiltrate' OR 'edema' | No ML model. Rule-based ALI Sniffer based on presence of PaO2/FiO2 < 300 and radiology report features | N/A | N/A | Sensitivity 96.3% (93.6 - 98.1); Specificity 89.4 % (88.4 - 90.4); PPV 46; NPV 99.6 |

Our goal in this study was to build a tool capable of identifying ARDS in a manner similar to real-time clinical evaluation in patients with severe hypoxemia (PaO_2_:FiO_2_ ratio < 150) receiving mechanical ventilation. To increase the likelihood that the algorithm developed would be applicable to a real-world patient population, we focused on building/refining our model using data collected in ICUs of one of the most diverse hospital networks in the United States (Northwell Health, New York), and focused on data readily available to clinicians within a 24-hour window preceding the diagnosis. After feature extraction from medical records that included laboratory values and natural language processing (NLP) of radiological reports, we implemented a unique, iteration based framework that allowed us to build an effective discriminator that can potentially be implemented in clinical sites to inform on ARDS diagnosis in real time. In doing so, we identified clinical parameters capable of augmenting real-time clinical assessment of ARDS in the ICU.

## METHODS

### Dataset

As our goal in this investigation was to construct a discriminative model for ARDS among patients requiring mechanical ventilation, it was critical to construct cohorts of patients with physician-diagnosed ARDS versus respiratory injury from another source (herein ARDS and non-ARDS, respectively; see **Figure 1**). Our analysis used a dataset of 1,263 mechanically ventilated patients with severe hypoxemia (PaO_2_/FiO_2_ < 150) admitted between May-2016 and April-2019 to the ICU in 12 hospitals at Northwell Health in New York State. Northwell Health is New York State’s largest healthcare provider and cares for a socio-economically and racial/ethnically diverse patient population. The screening period took place during the flu seasons (defined as October through April of each year) starting October 2016 through April 2019. The dataset was built using an algorithm to identify all invasively mechanically ventilated patients who could potentially have severe ARDS using the following query steps: 1. All intubated patients; 2. Admitted to an intensive care unit (ICU); 3. PaO_2_:FiO_2_ ratio of < = 150 based on arterial blood gas results, on at least two consecutive samples; 4. PEEP > 5cm H_2_0, and ; 5. Age >= 18. For patients identified from this query, radiology reports most proximal to the timing of the PaO_2_:FiO_2_ criteria were extracted into a relational database (REDCap) and included an indication (Y/N) of whether they carried an ICD code for respiratory failure (using codes J80.X, J96.X, 518.81, and 518.82). These were not filters, but helped with the further refinement of the query as described below.

**Figure 1.**
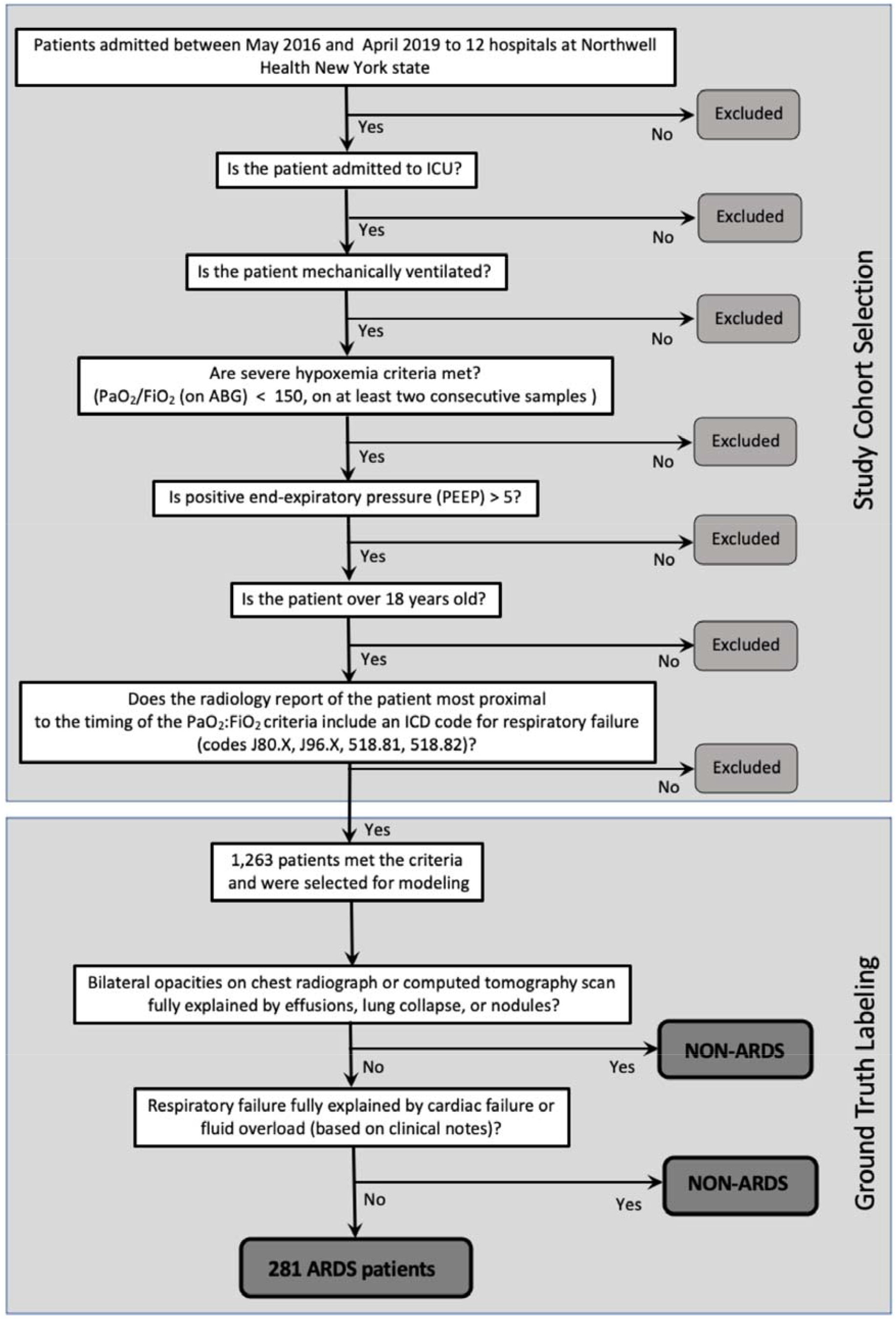
Flow diagram of study cohort selection and ground truth labeling by senior pulmonary critical care physicians. These criteria were used to define 281 gold standard ARDS patients from among the patients seen at Northwell Health between May 2016 and April 2019.

This process occurred prospectively to search for new patients, and records of those already selected on previous days were updated with newly available data. Next, two additional criteria described in more detail below had to be satisfied for the presence of moderate to severe ARDS: a) bilateral opacities on chest radiograph or computed tomography scan not fully explained by effusions, lung collapse, or nodules, and b) respiratory failure not fully explained by cardiac failure or fluid overload (based on clinical notes). All data including demographics, lab and image results, flow sheets, medications, and notes, were exported to an SQL database for further data analysis. All patients who met the screening criteria of potentially having severe ARDS were auto-identified in the EMR system as described above and filtered into a relational database (REDCap) with pre-defined fields of characteristics important for ARDS diagnosis. This included radiographic reports and clinical notes at the time of meeting severe hypoxemia inclusion criteria. Clinicians had access to the full electronic health record to further investigate the patient’s clinical course as needed and the auto-detection algorithm was iteratively updated for other features important to include in the database after observations of clinician interaction with the patients’ electronic health record files.

### Clinical Evaluation

For the identification of true positive severe ARDS, three pulmonary critical care physicians independently reviewed the records in the REDCap database and categorized patients into those with clinically confirmed ARDS and other diagnoses of respiratory failure. A senior pulmonary critical care physician reviewed a random sample of 20% of all confirmed ARDS cases.

### Automated Feature Extraction from Radiological Reports

Next, we examined unstructured free text observations from chest X-ray and CT scan reports for the entire cohort of mechanically ventilated patients with respiratory failure. Minimal preprocessing was applied to the notes before conversion to numerical feature vectors. Specifically, a manual spelling correction was applied, the stop words were removed, and a lemmatization was performed using the NLTK python package^14^. The notes were then summarized to contain only the sentences with keywords manually curated based on relevance to bilateral infiltration and pulmonary edema. The subset of relevant keywords was based on an initial list extracted from all available radiology reports^15^, and refined by the clinical team based on their domain knowledge (see **Table 2** for the list of keywords).

**Table 2.** List of keywords used for text summarization from radiographic reports, organized by domain (ARDS/Cardiovascular/Other).

|  |  |
| --- | --- |
| **ARDS key words** | ARDS, biapical, bibasilar, bilateral, edema, infiltrate, parahilar, perihilar, |
| **Cardiovascular-related keywords** | cardiac, cardiomegaly, cardiac, congestive, enlarged, failure, heart, flow |
| **Other key words** | widespread, blood, coarse, consolidation, cysts, extensive, brochogram, bronchial Lung, opacification, groundglass, opacities, opacity, pulmonary, peribronchial cuffing, pneumonia, pneumocyte, air, airspace, alveolar, aspiration, bronchovascular, bullae, air, airspace, coalescent, diffuse, dilatation, pleural, effusion, fibrosis, inverted, septal, lines, multifocal, pattern, pedicle, proliferation, reticular, size, vascular, volume |

Radiology reports were then converted into numerical feature vectors using doc2vec^16^, implemented using the *spacy* package for Python. These numerical word embeddings inherently contain the relationships between words, and thus are capable of retaining the relational structure of the text. We removed numbers and non-alphanumeric characters and stop words to prepare the summarized texts for doc2vec. The data were then randomly split into training and test sets, and a Random Forest model trained with performance measured using the (withheld) test set to determine discrimination between ARDS and non-ARDS. To have a reliable estimate of performance, we used a cross-validation framework in which the process was repeated 100 times with different training and test sets in each iteration. The final performance metric is the median of the metrics acquired for the 100 iterations.

### Medical Lab Test Data Processing

The dataset included the results of all lab tests performed during patient admissions. In total, the dataset consisted of 1933 distinct lab tests, for 1,263 total patients. Lab tests with >50% missingness were excluded from the analysis (**Table S1)**.

Timing of ARDS onset was based on the first PaO_2_/FiO_2_ measurement that met inclusion criteria. To ensure that the model would only contain information available to clinicians within 24hrs of ARDS onset, only lab tests taken prior to ARDS onset were considered as features in the laboratory analysis. Lab values considering two timepoints were included: static measurements in the 24 hours prior to PaO_2_/FiO_2_ inclusion criteria, and the rate of change (slope) of continuous lab test values over the prior four days. The slope for rate of change in labs was measured by fitting a linear regression model on the data. If a measurement was missing for a specific day, we used the last observation carried forward (LOCF) imputation approach used commonly in longitudinal studies. This method assumes that the value has stayed the same as the prior value. The absolute value of the slope represents the rate of change over time, and the sign (+/-) represents the direction of change (increased/decreased).

### Machine Learning Analysis

In addition to selection of an ML algorithm, ML workflows often involve selection of model parameters, feature reduction approaches, and feature normalization techniques. These are selected through combinations of known best practices, empirical observation, or performance scoring-based permutation. To allow for an unbiased determination of these model parameters and maximize the ability of the model to automatically adjust to additional datasets, we implemented an iterative processing framework in which data pre-processing parameters, ML models, and ML hyperparameters were simultaneously permuted to find an optimal configuration (**Figure S1**). Before beginning the learning process, the data were divided into training and validation sets of 80% and 20%, respectively, and all permutations were evaluated based on iterative bootstrapping of the training set. All model parameters mentioned below were identified through this process.

Classification models were built using features extracted from laboratory test results, features extracted from the radiological reports, all features combined, and outputs of combined models. This allowed for evaluation of the relative impact of these data sets to discriminate between ARDS versus non-ARDS. <u>In total we built five models</u>: model 1 included frequencies of each lab test’s acquisition (per hour) from the period of admission to the PaO_2_:FiO_2_ measurement time; model 2 included laboratory features (results) and mechanical ventilator parameters which together we term as ‘lab values’, and demographic information (i.e. age, gender) as well as comorbidities; model 3 included keywords from the radiology reports derived using the NLP method described above; model 4 used both model 2 and 3 inputs (i.e., both lab values and radiology reports), and; model 5 included the model outcomes from model 1, 2 and 3 combined in a logistic regression model.

For ARDS classification based on the lab tests (models 1 and 2), we used the Random Forest classifier implemented in the Python sklearn package^17^. The Random Forest (RF) model provides an importance score for each feature, and has hyper-parameters (e.g. number of trees) that can require tuning. To identify an optimal model for this dataset, we tuned: *max depth, min samples split, min samples leaf, max features, number of estimators*, and *criterion*, used a Bayesian approach implemented in the Optuna Python package^18^. To evaluate performance, we randomly split the dataset of 1,263 patients into training (80%), and final test sets (20%). The final test set was not used in any part of the analysis, and was considered an independent set to report the performance. The initial training set was then split randomly 100 times into training (70%) and validation (30%) sets. The RF hyper-parameters were tuned based on the performance on the validation set. The trained models in each iteration were stored to later create an ensemble model. We used the same approach for prediction of ARDS based on the radiology reports (model 3). Twenty percent of the data was considered a final test set, and the remaining data was split in a cross-validation framework (70% training, and 30% validation) 100 times to obtain a distribution of the model performance and to get a robust estimation of performance metrics. The trained models in each iteration were again stored to later create an ensemble model. This same process was again repeated for the model that included the combined features (i.e., radiology report features and medical lab results; model 4).

The performance of all models was measured as the area under the Receiver Operating Characteristic (ROC) curve (AUC); and the precision at 20% recall on the final test set. Note that precision at a given recall indicates the positive predictive value of a test at a given, fixed sensitivity. So, for example, precision at 20% recall is the equivalent of the positive predictive value at a sensitivity threshold in which at least 20% of the positive cases have been identified, and thus suggests the positive predictive value of the classifier for the most “identifiable” 20% of patients. Unlike AUC, this score ranges from 1.0 (perfect precision) to the ratio of true positives to total cases (the Prior), as this is the performance we would expect from a random classifier. It is being included as a metric here because precision/recall values are less prone to inflation due to class imbalance (e.g. more negatives than positives in the population) than AUC.

The analysis with combined features (model 4) was based on the existence of interaction between lab result features and features extracted from radiology notes. The improvement in performance upon considering this interaction might be offset by increasing the number of features and subsequently the estimation variance. As an alternative approach, in model 5 we used the outputs from models 1, 2, and 3 (RF models), as inputs to a logistic regression model to estimate the data label (i.e. ARDS, or non-ARDS). Similar to the previous modeling approaches, the training was performed in a cross-validation framework with 100 random splits of data into training and test sets. Hyperparameter optimization was determined through permutations of each model (**Table 3**) with average precision score as the performance metric.

**Table 3.**
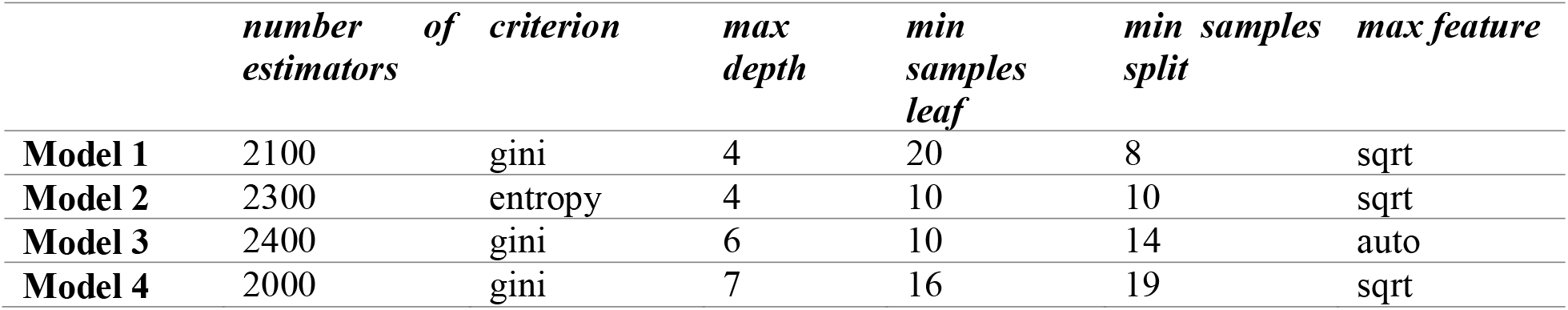
Hyperparameters for models 1, 2, 3 and 4 that provided the highest performance (avg precision score).

An RF model consists of a number of decision trees, and every node in the decision tree is conditioned on a single feature. As a result, the heterogeneity of samples in each set (a.k.a. impurity) can be reflected by the change in Entropy of the samples when a particular feature is used within a node of a decision tree. This average Entropy change was interpreted as feature importance, wherein the features that more greatly decrease the impurity get a higher importance score. However, the importance score is a *relative* measure and can only be compared within a specific training setup (specific feature set, and model hyper-parameters). Therefore, a permutation-based test was used to identify when features were significantly contributing to model construction. Specifically, target labels were randomly permuted at least 1000 times, and the distribution of importance score for each feature score was measured. The empirical p-value was determined as the number of times (out of the number of total permutations) that a feature had a higher feature importance in a random-model compared to its feature importance in the true model (as previously described^19^). Those features with p<0.05 were interpreted to be significantly contributing to the model.

## RESULTS

A total of 1,263 patients were prospectively identified as potentially meeting the Berlin criteria of severe ARDS based on PaO_2_:FiO_2_ levels, among mechanically ventilated patients at Northwell Health hospitals during the flu seasons of 2016 - 2019. Manual screening identified 293 of these patients as true positive severe ARDS cases which were confirmed by a senior clinician. The remaining 1,091 were determined to be false positives, and were then further classified into 11 sub-categories: Pneumonia (unilateral), Pulmonary Edema (Cardiogenic or Neurogenic), Atelectasis, Chronic Lung Disease/ILD COPD/Asthma, Pneumothorax, Pleural Effusion, Pulmonary Embolism, Pulmonary AVM, Intra-cardiac shunt, Pulmonary Contusion, Pulmonary Hemorrhage (**Table S1**).

There was a modest, but significant age difference between ARDS and non-ARDS groups (median age of 63 for the ARDS group versus 67 for non-ARDS, p-value=0.014, Mann-Whitney Test; **Table 4**). The distributions of age were found to be similar, however, there was an additional peak between ages 30-40 for ARDS patients. Race appears equally distributed, and in both groups males comprise a greater proportion of patients than females.

**Table 4.**
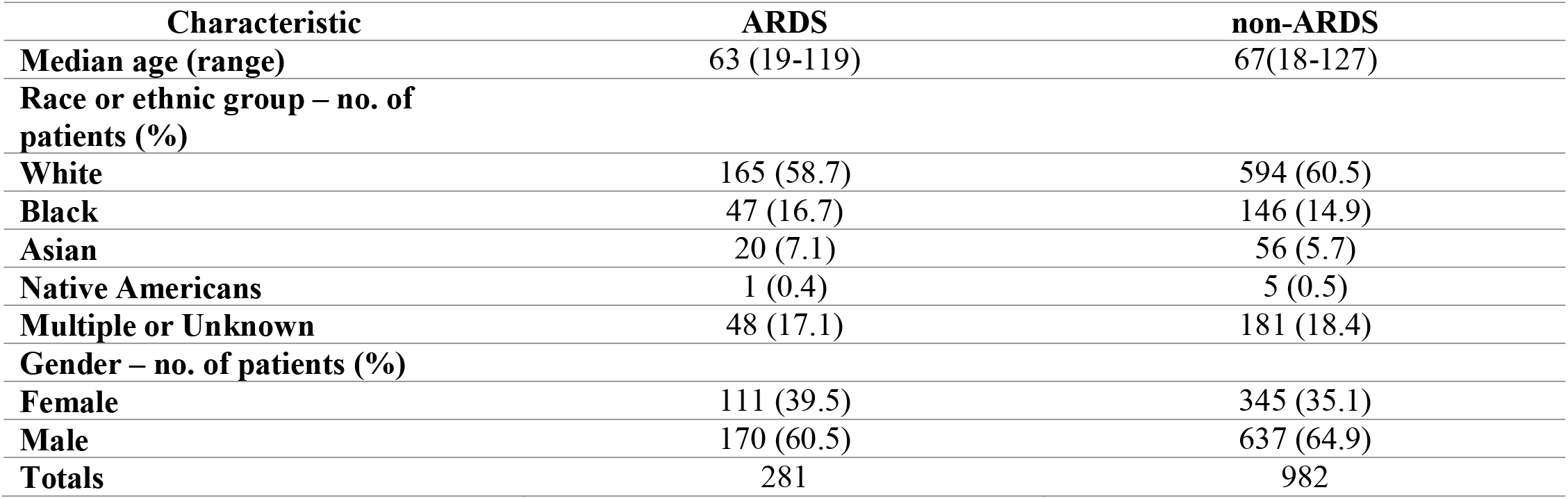
Demographics for ARDS and non-ARDS patients in the cohort.

| **Characteristic** | **ARDS** | **non-ARDS** |
| --- | --- | --- |
| **Median age (range)** | 63 (19-119) | 67(18-127) |
| **Race or ethnic group – no. of patients (%)** |  |  |
| **White** | 165 (58.7) | 594 (60.5) |
| **Black** | 47 (16.7) | 146 (14.9) |
| **Asian** | 20 (7.1) | 56 (5.7) |
| **Native Americans** | 1 (0.4) | 5 (0.5) |
| **Multiple or Unknown** | 48 (17.1) | 181 (18.4) |
| **Gender – no. of patients (%)** |  |  |
| **Female** | 111 (39.5) | 345 (35.1) |
| **Male** | 170 (60.5) | 637 (64.9) |
| **Totals** | 281 | 982 |

We constructed several classification models based on independent feature sets to allow us to objectively determine the predictive capacity of each in identifying ARDS among mechanically ventilated patients (**Figure S1**). When examining the data, we observed that patients had a disparity in the availability of laboratory parameters and wanted to determine whether the presence of an order for a lab test by itself was informative. Therefore, in the first model (Model 1) we used only the frequencies of lab test acquisition as features (i.e., not the lab values themselves). This model achieved fair performance (Area under the Receiver Operating Characteristic Curve (AUC) 0.71 (95% CI [0.68, 0.73]), Precision at 20% Recall (P@20R) 0.5; **Figure 2**). This confirms that there is discriminative information content in the frequency of tests performed.

**Figure 2.**
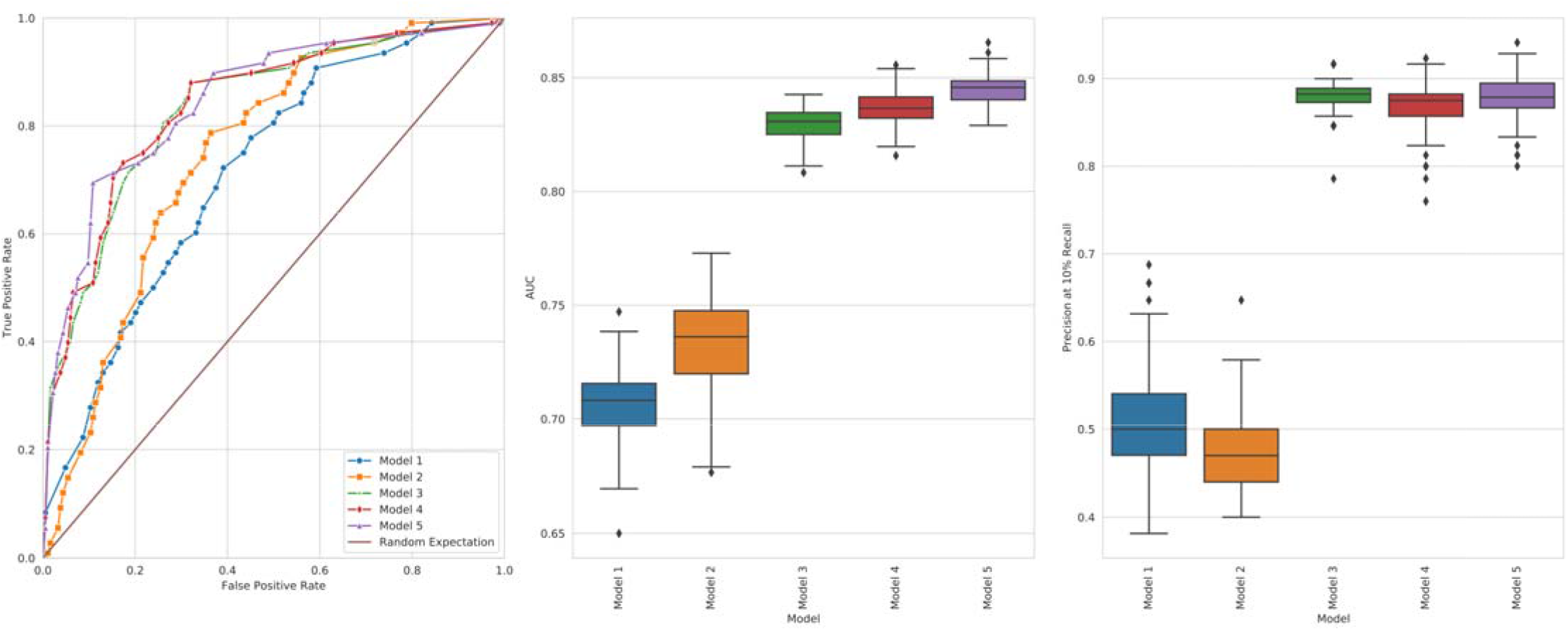
Classifier performance. Five sets of models were trained using features extracted from (1) frequency of the 1933 medical lab tests (per hour) from the admission to P/F measurement; (2) medical lab tests, mechanical ventilation measurements, demographic information; (3) radiology reports (keywords in **Table 2**); (4) combined medical lab tests with radiology reports features; (5) model (1), (2) and (3) outputs (two stage model). The training was performed in a cross-validation framework with 100 splits, and the performance metrics were measured for each split. Left: The receiver operating curve of the trained models. Middle: The area under the ROC curve; and Right: the precision (at 20% recall) for the same models.

Next, we created a model using laboratory values themselves in addition to the demographics and comorbidities (Model 2; **Figure 2**). We eliminated lab features that had >50% missingness in the cohort, so as to focus on construction of a model potentially generalizable across multiple clinical cohorts. In other words, missingness for a dataset like ours implies that the clinician did not believe the feature (lab value) needed to be measured. Performance prediction metrics showed a small increase in performance over Model 1 (AUC 0.75 with 95% CI [0.70, 0.76], 0.54 P@20R with 95% CI [0.42, 0.54]).

In Model 3 we examined ARDS versus non-ARDS classification using only RF models trained on features extracted from radiology reports. Performance of this model was higher than that seen in the laboratory test model (AUC 0.79 with 95% CI [0.82, 0.84], P@20R 0.73 with 95% CI [0.85, 0.9]; **Figure 2**). Model 4 included features used in both Models 2 and 3, and again, classification performance was found to incrementally improve (AUC=0.8 with 95% CI [0.82, 0.85], P@20R=0.74 with 95% CI [0.81, 0.92]). Finally, Model 5 provided the highest classification accuracy (AUC=0.85 with 95% CI [0.83, 0.86], P@20R=0.89 with 95% CI [0.83, 0.92]) by combining the outputs of the models 1, 2 and 3 in a logistic regression framework.

With the models constructed, we next applied a permutation-based method to investigate features that were contributing to classification accuracy. We performed this analysis for Models 1, 2 and 3 only, as these models contained all features included in subsequent models. First, in Model 1 (**Figure 3**), the frequency of myelocytes and metamyelocytes seen in lab reports, as well as a higher frequency of blood gas measurements rank as important for identifying ARDS among mechanically ventilated patients. For Model 2 (**Figure 4**), arterial pH, arterial oxygen saturation, serum albumin, and total serum calcium results were ranked the most relevant static lab results in the 24 hours prior to ARDS diagnosis. The following features were most relevant in predicting ARDS based on change over time in the 4 days prior to ARDS diagnosis (**Figure 4b**): arterial partial pressure of oxygen (PaO_2_), arterial pH, and eGFR. For Model 3, the features were acquired from a non-invertible word2vec transform on the text that obfuscates the relation between features and the text. Therefore, the feature importance is not meaningful for discrimination between ARDS and non-ARDS reports.

**Figure 3.**
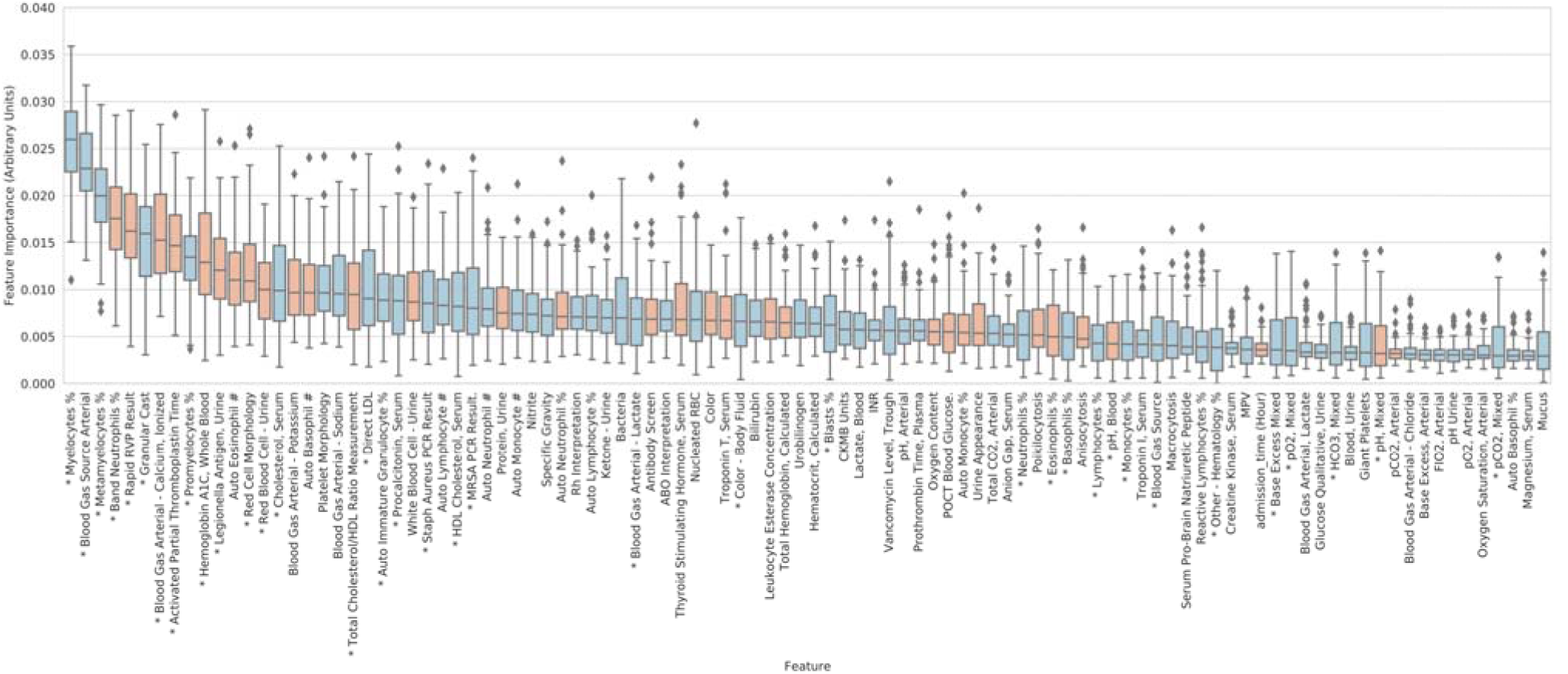
Feature importance scores for Model 1. Feature importance scores for a random forest classifier trained using frequency of timing of laboratory tests (top 100 features shown). The orange (or blue) color for a box indicates that the associated lab test was acquired in a higher rate in the ARDS patients (or non-ARDS patients). For example, Myelocytes % acquisition rate is a feature with high importance in discriminating between ARDS versus non-ARDS, and is higher in ARDS compared to non-ARDS.

**Figure 4.**
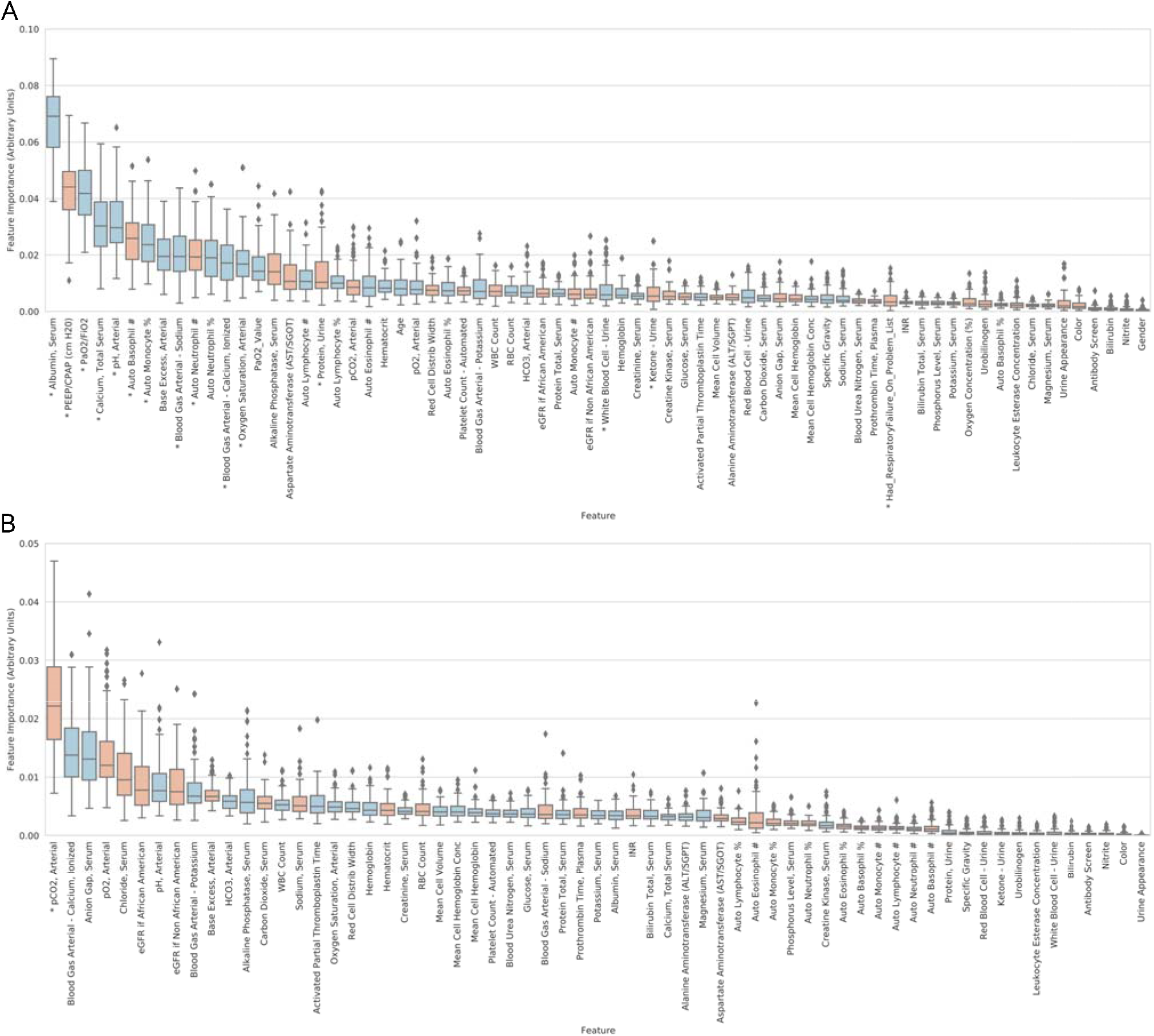
Feature importance scores for Model 2. **(A)** The feature importance scores for Model 2 using absolute values extracted using a random forest model in a cross-validation framework. The orange (or blue) color for a box indicates that the associated lab test has a higher value in the ARDS patients (or non-ARDS patients). For exa ple, PH Arterial is a feature with high importance in discriminating between ARDS versus non-ARDS, and is lower in ARDS compared to non-ARDS; and alkaline phosphatase is higher in ARDS compared to non-ARDS. (**B**) The feature importance score for model 2 using **change in slope** of the values extracted using random forest model in a cross-validation framework. The orange (or blue) color for a box indicates that the magnitude of change in the associated lab test is greater in the ARDS patients (or non-ARDS patients). For example, change in arterial PCO2 has high importance for discriminating between respiratory failure likely due to ARDS versus non-ARDS, and a greater magnitude of change in arterial PCO2 over four days is more likely to be seen in ARDS as compared to non-ARDS; similarly, change in anion gap has high importance for discriminating between respiratory failure likely due to ARDS versus non-ARDS, and a greater magnitude of change in anion gap is more likely to be seen in non-ARDS as compared to ARDS. The asterisk below each feature indicates its importance score has passed the significance threshold (p<0.01).

Our clinical evaluators used both radiological reports and clinical notes for their retrospective assessment of ARDS (see **Methods**), consistent with the Berlin criteria. However, we saw fair performance for models 2 (reports only) and 3 (lab tests only), which prompted us to examine whether subsets of patients could be accurately classified using either data type alone. To this end, we examined the probabilities of classification for individual patients (whether the model suggests classification into ARDS versus non-ARDS groups) from models trained using these datasets (models 2 and 3; **Figure 5**). There was a moderate correlation of the classification probability scores for each patient (r=0.4, p<0.01), suggesting a non-random but modest agreement between classification scores generated by models trained on laboratory tests and radiological reports alone. In other words, consistent with what is seen clinically, while both laboratory tests alone and radiological reports may be indicative of ARDS, there may be key implicators only contained in one data type for a given patient, thus explaining the improved performance of classifiers using both datasets.

**Figure 5.**
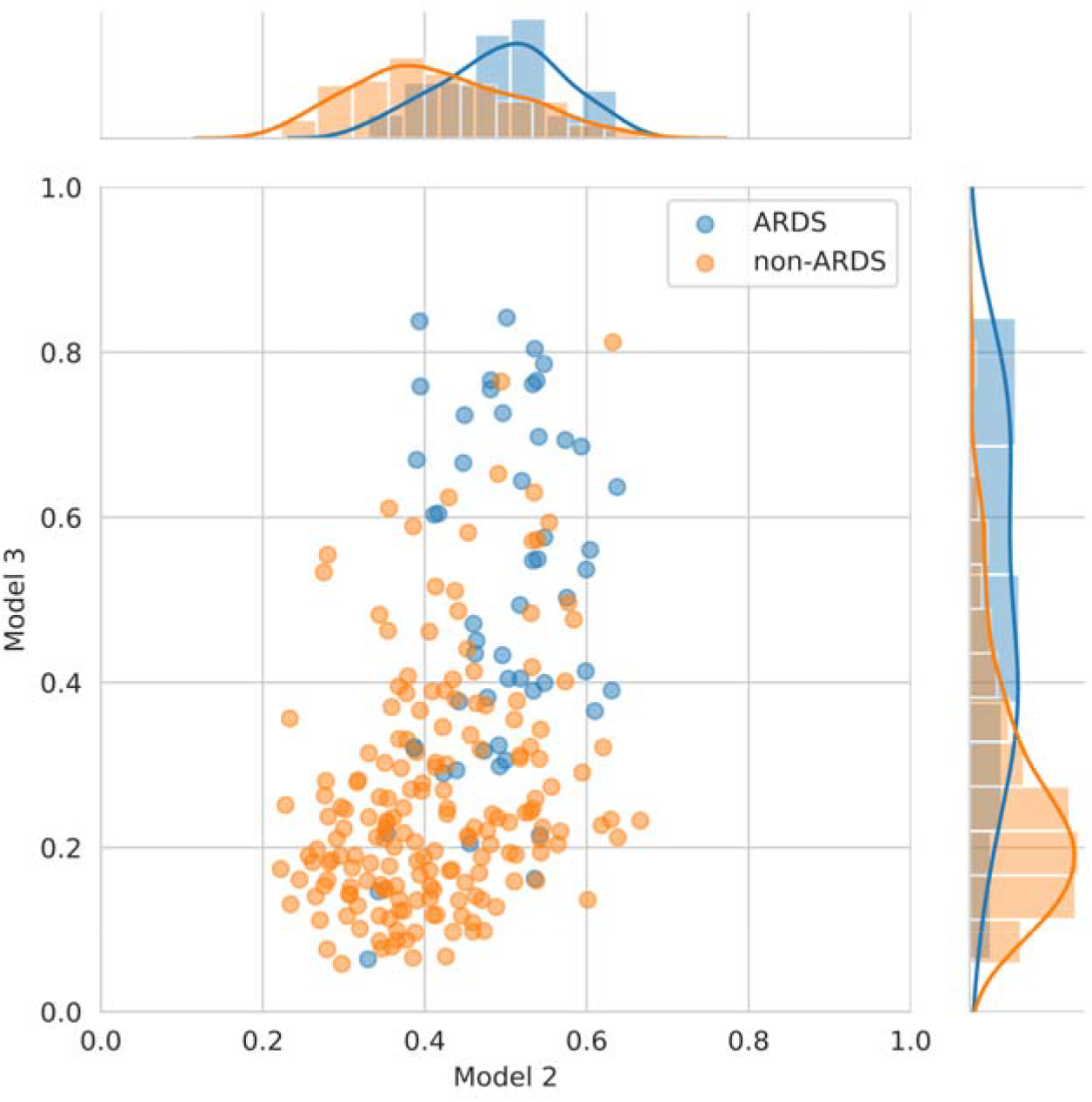
Performance by patient for Models 2 and 3. The scatter plot shows the output of Model 3 (trained using radiology reports) versus Model 2 (trained using laboratory tests) for both ARDS (orange) and non-ARDS (blue) patients. The distributions of the probabilities for Models (2) and (3) are shown on upper and right side of the scatter plot for both ARDS and non-ARDS patients. Underlying heterogeneity among patients makes a definitive classification difficult even with access to both radiological reports and laboratory tests.

Next, we examined patients that were incorrectly classified by our most accurate model to determine if there were any consistent features among patients that could not be classified by our framework. Specifically, we measured the classification error within the ARDS group (i.e., those wrongly classified as ARDS), and we explored the relationship between the error and mechanical ventilation parameters and demographic information using PLS regression (**Figure 6**). Based on the result from our best model (i.e., Model 5), our machine learning framework has a higher error rate for older patients and performs better for patients with a history of respiratory failure. This suggests that ARDS risks outputted from this model for older patients, specifically those with no previous history of respiratory failure, should be interpreted with caution before therapeutic action.

**Figure 6.**
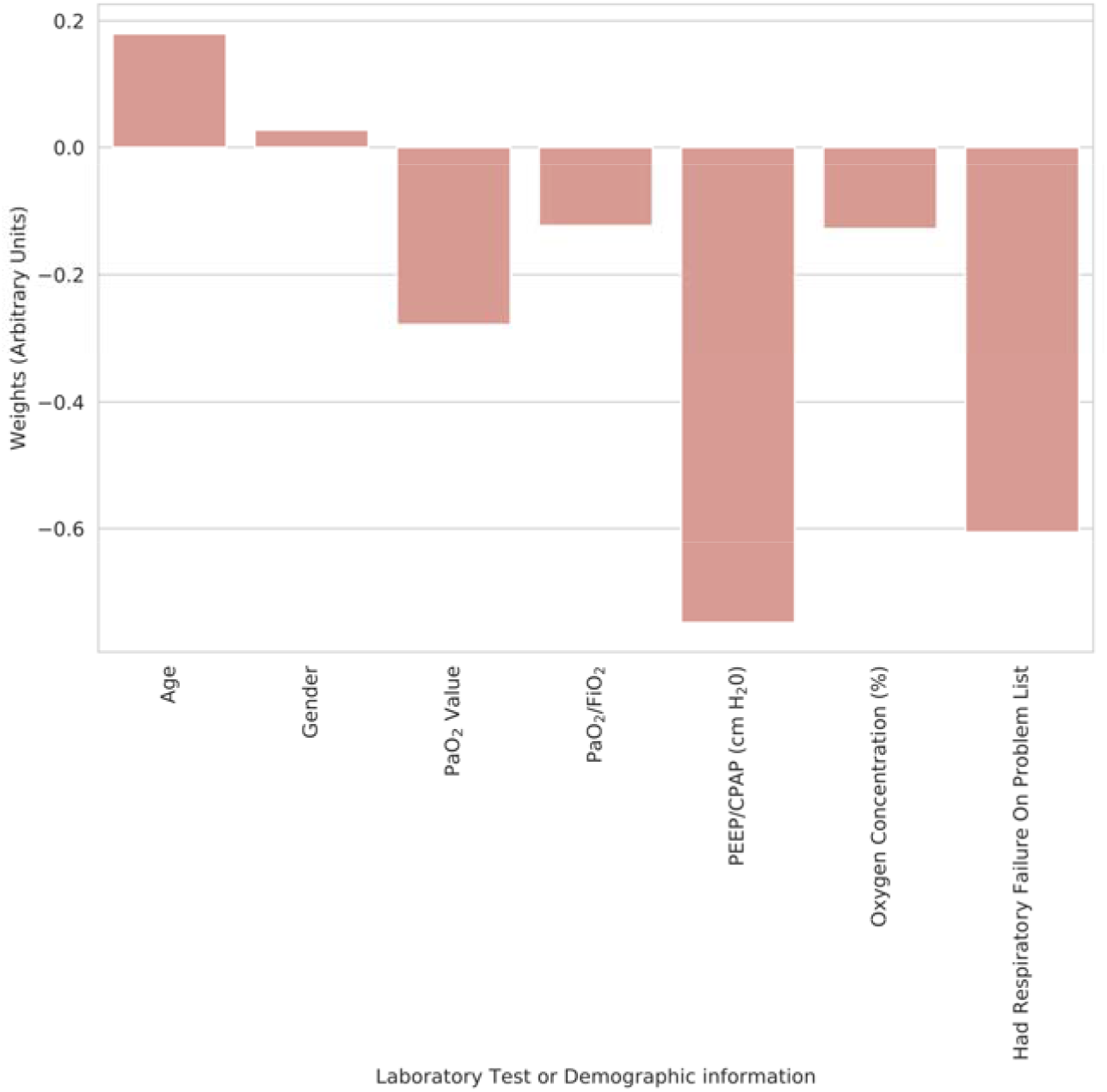
Classification error by feature. The relationship between classification error, ventilation, and demographic information (x-axis) plotted against error correlation (arbitrary units; y-axis). The predictions generated by Model (5) had a higher error with increasing age of the patient, and error was inversely correlated with respiration failure history, meaning the framework performed better for patients with a history of respiratory failure.

Finally, we sought to determine if the models could be used to identify features unique among ARDS patients expiring as a result of their illness, versus non-ARDS patients. Specifically, we re-trained the above models to predict mortality among either ARDS or non-ARDS patients, with the expectation that the resulting feature importance analysis would identify features predictive of mortality uniquely in ARDS. We found that a number of features (notably pH) were more strongly indicative of mortality for ARDS patients (**Figure S2a**), as compared to non-ARDS patients (**Figure S2b**).

## DISCUSSION

Discrimination of ARDS from other causes of respiratory failure requiring mechanical ventilation remains a challenge despite extensive study. The propensity of the Berlin criteria to produce substantial false-positives, coupled with the lack of available biomarkers for ARDS has necessitated manual review of radiology reports and laboratory tests by physicians, which can introduce subjectivity. This subjectivity not only impacts patient care, but introduces inconsistency in clinical trial inclusion for ARDS. Under-diagnosis of ARDS has been found by observational studies^20^, and even within the context of a randomized controlled clinical trial misclassification if radiology reports has been demonstrated^21^. Consequently, a machine-learning based assessment framework would: 1) provide physicians with a real-time assessment of ARDS likelihood, 2) allow for standardized ARDS assessment across ICUs, and 3) identify novel clinical determinants specific to ARDS for further study. Importantly, the machine-learning framework must emulate objective clinician practice with readily available data, in this case laboratory tests, radiology reports, and mechanical ventilator features.

The model building process we employed independently optimized five model configurations, each leveraging different input feature sets. By comparing these respective models we were able to more objectively determine the relative contribution of each of these feature sets to classification performance. As a baseline model, and to examine the importance of measurement frequency on classification, we began by including only features corresponding to the frequency of measurement of laboratory tests. This produced a surprisingly robust classifier, suggesting that patterns in clinical care, including ordering of tests, is itself predictive of patient outcome. This is an important observation, and suggests that alterations in measurement procedure, even owing to altered and/or updated risk assessments of ARDS, should be accounted for in future models.

Next, we examined the contribution of laboratory test values and radiological reports, both apart and together, in discriminating ARDS among our ventilated patients. Unsurprisingly, a model leveraging both of these datasets in combination outperformed models built using either data source alone, mimicking the diagnostic process of a clinician. The high performance of our final model (Model 5) suggests that all three components - frequency of test measurement, laboratory test results <u>including changes over time</u>, and radiological reports – improve model discrimination.

To place our study in context with the work of other colleagues who have endeavoured to identify ARDS we summarize these in **Table 1**. Most of the studies have been focused on prediction the onset of ARDS or Acute Lung Injury (ALI) which in prior versions of accepted categorization encompassed ARDS as a more severe form of lung injury. In contrast, our study was aimed at discriminating cause of respiratory failure requiring invasive mechanical ventilation to be ARDS versus another cause. The study most closely resembling ours is the recent study by Le et al.^22^ which similarly attempts to detect ARDS although their study did not include radiography features and did not include changes in features over time. The AUC for the model was similar to ours (AUC 0.83) and the most influential features - albeit only provided for the non-mechanically ventilated cohort - included antibiotics, vital signs, and pH. Our study did not include treatments or vital signs but similarly found pH to be a highly-ranked feature for discriminating between ARDS and non-ARDS causes of respiratory failure. Other models which have looked at ARDS prediction have found vital signs, in particular heart rate, mean arterial pressure and respiratory rate to be important for predicting ARDS. This is likely due to the overlap between sepsis and ARDS. Our models did not include treatments or vital sign information. Laboratory features of importance that have been found by other investigators include low hematocrit, low glucose, low sodium, normal platelet count, and elevated white blood to increase the risk of ARDS. Laboratory features of importance in our models for the 24 hours prior to meeting potential ARDS criteria included arterial blood gas values (lower values of pH and PaO_2_, and higher PaCO_2_ for ARDS classification), and lower values for serum albumin, calcium and sodium predicting ARDS rather than non-ARDS causes of respiratory failure. Changes in laboratory values that were important included arterial blood gas values (greater magnitude of change in PaO_2_, PaCO_2_, and lower magnitude of change in PH for ARDS classification), anion gap (lower magnitude of change, ARDS), as well as eGFR and Prothrombin Time (greater magnitude of change, ARDS). Together, these features make clinical sense to be associated with ARDS more than with non-ARDS and to therefore help discriminate the two for mechanically ventilated patients. This can assist with the detection of ARDS shortly after mechanical ventilation is required and can support rapid implementation of ARDS treatment protocols, clinical trial enrollment and quality improvement monitoring. Earlier identification of ARDS could also assist with planning for potential transfers to tertiary care facilities with ARDS management experience.

The importance of discriminating ARDS from other causes of acute respiratory failure that require mechanical ventilation is underscored by our observation of differential risk factors for mortality. This could be important for longitudinal re-assessment of risk based on changes in these features, which in turn could inform prognostic conversations. These risk factors could also prompt causal pathway studies and discovery of treatment targets.

The most important limitation of this study is similar for all studies seeking to predict ARDS, which is the method by which true positive cases of ARDS are determined. Several studies have used an automated version of the Berlin criteria. This raises concerns about the validity of the true positive cases. Our automated query based on P:F values, bilaterality of infiltrates and invasive mechanical ventilation identified several false positive cases. We therefore assembled a group of physicians with ARDS experience who classified each of the potential cases. A senior clinician further confirmed a random selection of these cases which were coded by the clinicians. Nevertheless, subjective misclassification may still have occurred. Without a biomarker for true positive determination this remains the best method available currently. This underlines the importance of model validation prospectively. It should be noted that false negatives were not formally determined, as this would have required the manual check of all mechanically ventilated patients, and patients who were not intubated perhaps due to less severe illness of decisions to provide hospice care. However, it would be unlikely that an intubated ARDS case would have been missed by an algorithm based on P:F ratio such as ours.

Our goal was to determine if a trained machine learning classifier could accurately discriminate ARDS among other causes of respiratory failure requiring mechanical ventilation. Although our results suggest a useful model to augment clinician decision making, a fundamental next step will be prospective validation of model performance. While this model may begin to be evaluated to assess risk prospectively among patients in the Northwell Health, it is inevitable that some amount of refinement will have to occur with this or any ML model before generalization toward a more objective classification scheme at subsequent clinical sites. If we are successful in prospective validation, we must next determine whether it can be successfully implemented within clinical workflows to aid physicians in a difficult and potentially subjective definition that could impact patient care. This is particularly important for hospitals which do not have dedicated intensive care physicians who have extensive experience in ARDS diagnosis and management.

In summary, we have developed ML models which can assist with discriminating between mechanically ventilated patients who have ARDS vs, non-ARDS causes of respiratory failure, based on changes in laboratory features and radiology reports. Each of these features can be readily extracted from electronic health records to auto-populate models triggered by the start of mechanical ventilation.

## Data Availability

De-identified data will be made available via a Data Use Agreement (DUA) upon reasonable request

## Funding

Philanthropic funds to the Feinstein Center for Health Innovations and Outcomes Research

## SUPPLEMENT

**Table S1.** Medical lab tests, demographic information and ventilator measurements used for identification of ARDS

|  |
| --- |
| Potassium, Serum |
| Sodium, Serum |
| Blood Urea Nitrogen, Serum |
| Calcium, Total Serum |
| Creatinine, Serum |
| Carbon Dioxide, Serum |
| Glucose, Serum |
| Chloride, Serum |
| eGFR if Non African American |
| eGFR if African American |
| pCO<sub>2</sub>, Arterial |
| pO<sub>2</sub>, Arterial |
| HCO<sub>3</sub>, Arterial |
| Base Excess, Arterial |
| Oxygen Saturation, Arterial |
| pH, Arterial |
| RBC Count |
| Platelet Count - Automated |
| Hemoglobin |
| Hematocrit |
| WBC Count |
| Mean Cell Volume |
| Mean Cell Hemoglobin Conc |
| Mean Cell Hemoglobin |
| Red Cell Distrib Width |
| Anion Gap, Serum |
| Blood Gas Arterial - Potassium |
| Blood Gas Arterial - Sodium |
| Magnesium, Serum |
| Blood Gas Arterial - Calcium, Ionized |
| Phosphorus Level, Serum |
| Activated Partial Thromboplastin Time |
| Albumin, Serum |
| Bilirubin Total, Serum |
| Protein Total, Serum |
| Alanine Aminotransferase (ALT/SGPT) |
| Aspartate Aminotransferase (AST/SGOT) |
| Alkaline Phosphatase, Serum |
| INR |
| Prothrombin Time, Plasma |
| Auto Neutrophil % |
| Auto Lymphocyte % |
| Auto Monocyte % |
| Auto Eosinophil % |
| Auto Lymphocyte # |
| Auto Neutrophil # |
| Auto Monocyte # |
| Auto Basophil % |
| Auto Eosinophil # |
| Auto Basophil # |
| Creatine Kinase, Serum |
| Antibody Screen |
| Specific Gravity |
| Urine Appearance |
| Ketone - Urine |
| Nitrite |
| Color |
| Bilirubin |
| Protein, Urine |
| Urobilinogen |
| Leukocyte Esterase Concentration |
| White Blood Cell - Urine |
| Red Blood Cell - Urine |
| Age |
| Gender |
| PaO2 Value |
| PaO2/FiO2 |
| PEEP/CPAP (cm H2O) |
| Oxygen Concentration (%) |
| Respiratory Failure on Problem List |

**Table S2.** Features with high importance scores and magnitude of change, indicating ARDS as compared to non-ARDS as a cause of respiratory failure requiring mechanical ventilation.

| Features for which the value (in the case of labs, for the proceeding 24 hours before ARDS inclusion criteria) was statistically significant for discriminating between ARDS compared to non-ARDS | Median Importance Score | Value in ARDS group relative to non-ARDS |
| --- | --- | --- |
| PEEP/CPAP (cm H20) | 0.0397 | Higher |
| Albumin, Serum | 0.0346 | Lower |
| pH, Arterial | 0.0278 | Lower |
| Calcium, Total Serum | 0.0265 | Lower |
| Blood Gas Arterial - Calcium, Ionized | 0.0170 | Lower |
| PaO2/FiO2 | 0.0170 | Lower |
| pCO2, Arterial | 0.0167 | Higher |
| Base Excess, Arterial | 0.0157 | Lower |
| Oxygen Saturation, Arterial | 0.0152 | Lower |
| Blood Gas Arterial - Sodium | 0.0140 | Lower |
| Auto Basophil # | 0.0138 | Higher |
| Alkaline Phosphatase, Serum | 0.0130 | Higher |
| Age | 0.0129 | Lower |
| Auto Monocyte % | 0.0106 | Lower |
| PaO2_Value | 0.0099 | Lower |
| WBC Count | 0.0099 | Higher |
| Protein, Urine | 0.0087 | Higher |
| Auto Lymphocyte # | 0.0074 | Lower |
| Blood Gas Arterial - Potassium | 0.0073 | Lower |
| Auto Neutrophil # | 0.0061 | Higher |
| Auto Monocyte # | 0.0060 | Higher |
| Ketone - Urine | 0.0058 | Higher |
| Auto Eosinophil % | 0.0058 | Lower |
| Specific Gravity | 0.0038 | Lower |
| Urobilinogen | 0.0038 | Higher |
| Auto Eosinophil # | 0.0035 | Lower |
| Oxygen Concentration (%) | 0.0034 | Higher |
| Leukocyte Esterase Concentration | 0.0021 | Higher |
| Color | 0.0018 | Higher |
| Urine Appearance | 0.0015 | Higher |
| Antibody Screen | 0.0011 | Higher |
| Bilirubin | 0.0010 | Higher |
| Had_RespiratoryFailure_On_Problem_List | 0.0010 | Higher |
| Gender | 0.0006 | Higher |
| Nitrite | 0.0006 | Higher |

| Features for which the change in value over 4 days preceding ARDS inclusion criteria (slope) was statistically significant for discriminating between ARDS compared to non-ARDS | Median Importance Score | Relative rate of change (ARDS versus non-ARDS) |
| --- | --- | --- |
| * pCO<sub>2</sub>, Arterial | 0.0290 | Increased |
| * pO<sub>2</sub>, Arterial | 0.0236 | Increased |
| * pH, Arterial | 0.0130 | Decreased |
| * Anion Gap, Serum | 0.0102 | Decreased |
| * eGFR if African American | 0.0086 | Increased |
| * eGFR if Non African American | 0.0079 | Increased |
| * Prothrombin Time, Plasma | 0.0046 | Increased |
| Relative rate of change indicates the magnitude of the slope (change). For example, a greater change in arterial pCO<sub>2</sub> suggests ARDS classification instead of non-ARDS. |  |  |

**Figure S1.**
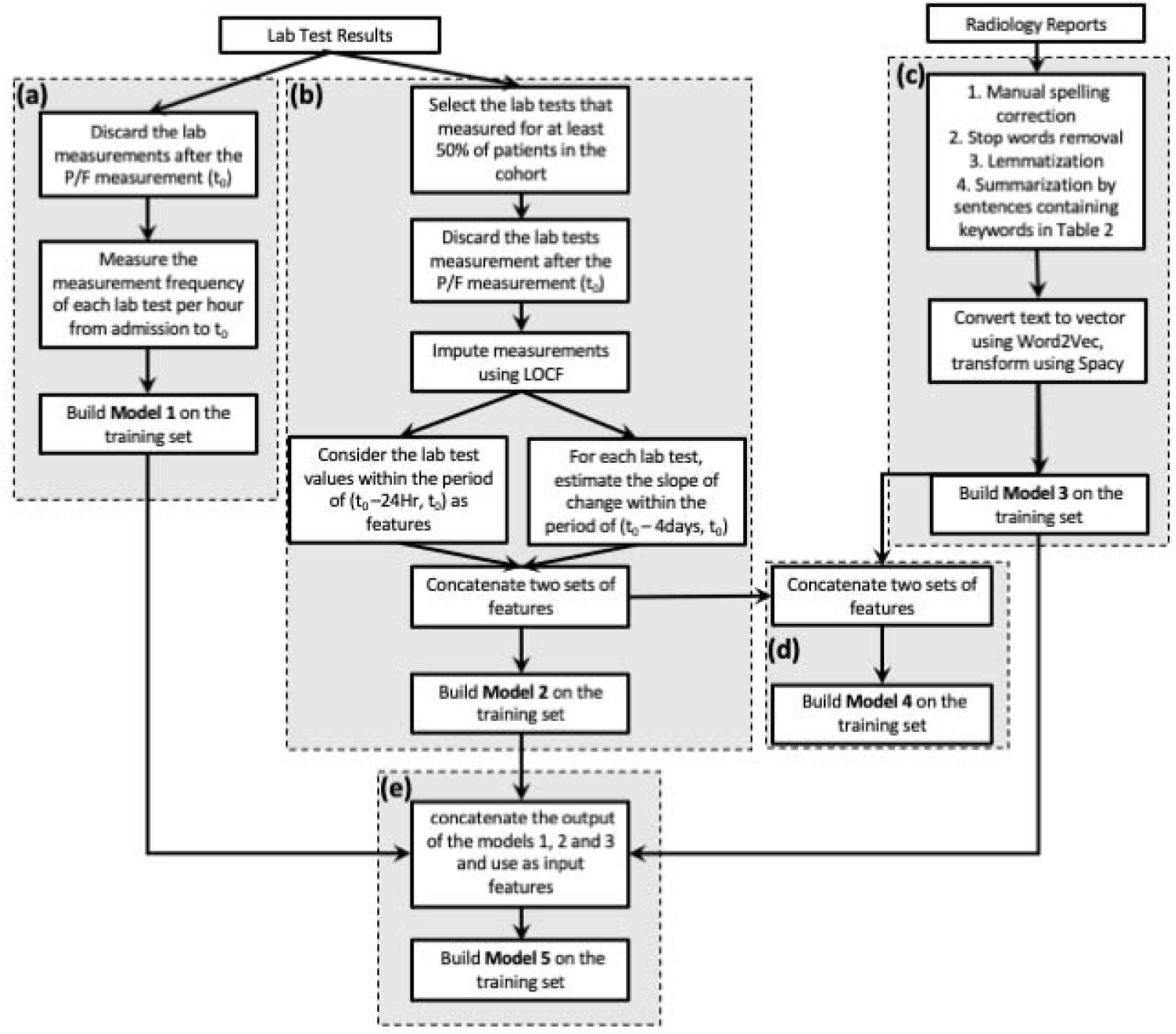
Overview of the Machine Learning Process. Multiple models were built leveraging different combinations of input features. In each case models were trained using an 80/20 training/test data split and evaluated over 100 permutations using test data. All five final models were evaluated using data withheld from the entire procedure.

**Figure S2.**
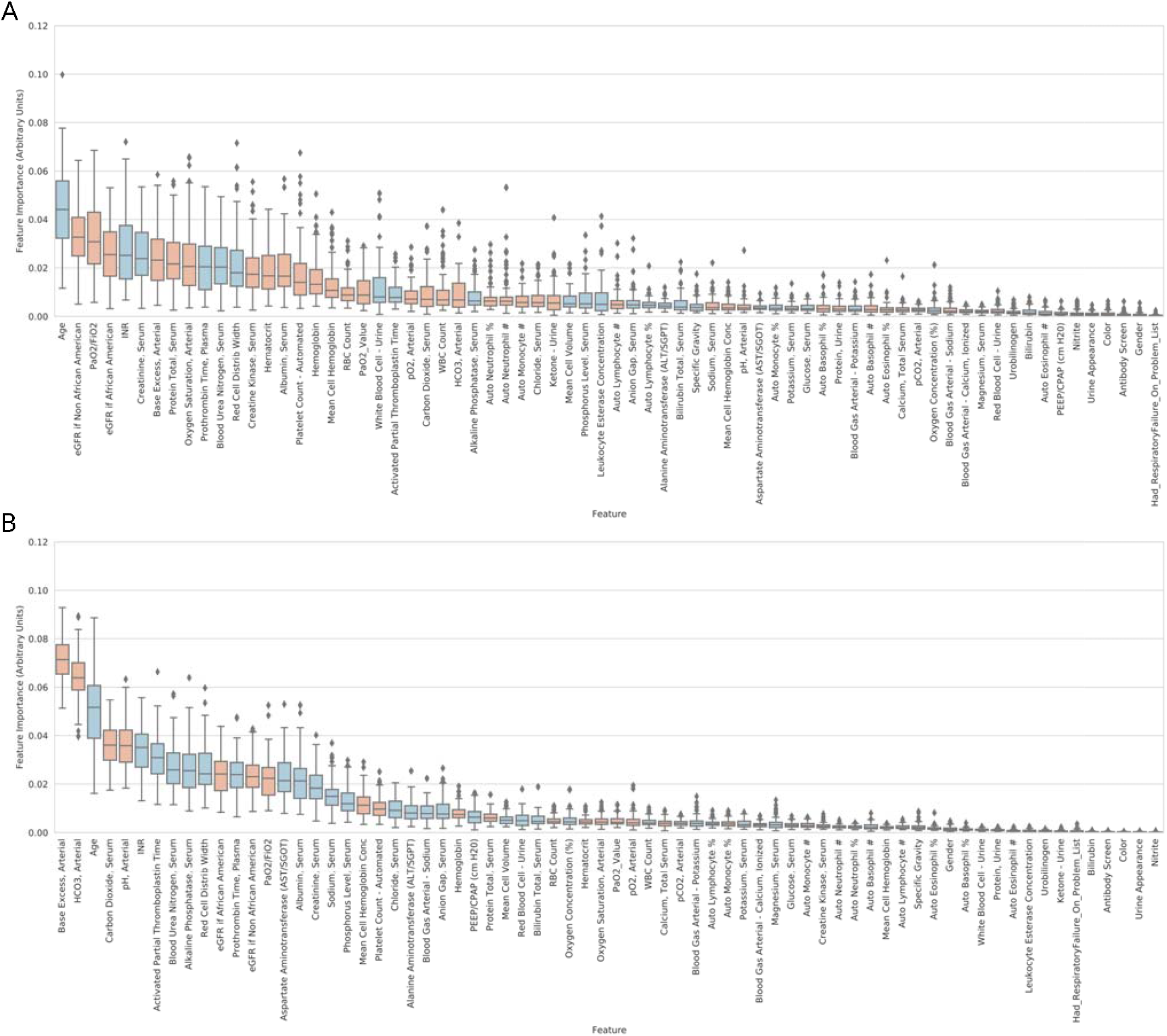
Feature importance scores for Mortality Prediction. **(A)** The feature importance scores for a prediction model discriminating ARDS patients who would not recover, versus those that were discharged (feature scoring and ranking performed as indicated above). Similarly, feature importance scoring is shown for patients that would not recover, versus those that would be discharged from the ICU for the non-ARDS group (B). The asterisk below each feature indicates its importance score has passed the significance threshold (p<0.01).

## Notes

### Competing Interest Statement

The authors have declared no competing interest.

### Funding Statement

This work was funded through philanthropic funds to the Feinstein Center for Health Innovations and Outcomes Research

### Author Declarations

Northwell Health / Feinstein Institute IRB #19-0598

